# Inpatient mortality and associated clinical factors among people living with HIV with cryptococcal meningitis in Uganda: a retrospective cohort study

**DOI:** 10.64898/2026.01.01.26343321

**Authors:** Joshua Kitimbo, Esther Buregyeya, Geofrey Mutole, John Paul Ibanda, Joseph Tumwine, Noah Kiwanuka, Thomas Buyinza

## Abstract

**Introduction:** Cryptococcal meningitis remains a leading cause of HIV-related mortality in sub-Saharan Africa despite expanded antiretroviral therapy coverage. Evidence on the burden of disease and inpatient mortality among people living with HIV (PLHIV) in routine care settings in Uganda remains limited. This study assessed the proportion of cryptococcal meningitis among HIV-related admissions and examined clinical factors associated with inpatient mortality.

**Methods:** We conducted a retrospective cohort study of adult PLHIV admitted with cryptococcal meningitis between January 2017 and December 2022 at a national referral hospital in Uganda. Diagnosis was based on cerebrospinal fluid cryptococcal antigen or India ink positivity. Data were abstracted from medical records and analysed using descriptive statistics and multivariable logistic regression to identify factors associated with inpatient mortality. Multiple imputation was used to address missing data.

**Results:** Of 3,042 HIV-related admissions, cryptococcal meningitis accounted for 21.4% (650/3,042). Medical records for 634 patients were analysed, among whom 39.3% (249/634) died during hospitalization. Factors independently associated with higher odds of inpatient mortality included convulsions, headache, vomiting, cryptococcal meningitis–associated immune reconstitution inflammatory syndrome, concurrent opportunistic infections, chronic kidney disease, anaemia, and severe immunosuppression (CD4 <200 cells/µL). Longer duration of hospitalization (≥7 days) and symptom duration of one to two weeks before admission were associated with lower odds of mortality.

**Conclusion:** Cryptococcal meningitis continues to account for a substantial proportion of HIV-related hospital admissions and inpatient deaths in Uganda. Mortality is associated with identifiable clinical and health-system factors, underscoring the need for early diagnosis, risk stratification, and optimized inpatient management for PLHIV with cryptococcal meningitis in resource-limited settings.

## Introduction

Cryptococcal meningitis (CM) is a life-threatening opportunistic fungal infection of the central nervous system that occurs predominantly among people living with HIV (PLHIV) with advanced immunosuppression. It is caused mainly by *Cryptococcus neoformans* and less frequently by *Cryptococcus gattii*, with infection typically acquired through inhalation of environmental spores followed by hematogenous dissemination to the brain, resulting in severe meningoencephalitis [1]. Without timely diagnosis and effective antifungal treatment, CM is associated with rapid clinical deterioration, neurological complications, and high short-term mortality [2, 3].

Despite substantial progress in HIV care and expanded access to antiretroviral therapy (ART), CM remains a major cause of HIV-related morbidity and mortality globally, with the greatest burden concentrated in sub-Saharan Africa (SSA) [4]. Of the estimated 38.4 million PLHIV worldwide, more than two-thirds reside in SSA [5], where late HIV diagnosis, advanced disease at presentation, and limited access to optimal management of opportunistic infections persist [6]. CM accounts for over 70% of cryptococcal disease among HIV-infected individuals and is responsible for approximately 20% of HIV-related deaths globally, highlighting its continued contribution to preventable mortality [7, 8].

In Uganda, CM is the leading cause of adult HIV-associated meningitis and remains a major contributor to inpatient morbidity, mortality, and long-term neurological disability among PLHIV [9]. Although national and international guidelines recommend routine cryptococcal antigen (CrAg) screening and standardized antifungal therapy for individuals with advanced HIV disease, implementation gaps remain common [7, 10]. Limited access to rapid diagnostics, inconsistent availability of amphotericin-based regimens, inadequate monitoring and management of raised intracranial pressure, and delayed care-seeking continue to undermine survival outcomes in routine hospital settings [3, 11].

Clinical factors such as severe immunosuppression, anaemia, seizures, concurrent opportunistic infections, renal dysfunction, and cryptococcal meningitis–associated immune reconstitution inflammatory syndrome (CM-IRIS) have consistently been associated with increased risk of inpatient mortality [12-14]. However, much of the existing evidence is derived from clinical trials, short-term cohorts, or mixed inpatient–outpatient populations, which may not adequately reflect routine care conditions in high-burden, resource-limited settings [4, 15]. In Uganda, contemporary evidence quantifying the burden of CM among HIV-related admissions and systematically examining inpatient mortality and its correlates over extended periods remains limited.

To address this gap, we analysed six years of routinely collected inpatient data from Kiruddu National Referral Hospital (KNRH) to quantify the proportion of cryptococcal meningitis among HIV-related admissions and to examine clinical and health-system factors associated with inpatient mortality. Generating such real-world evidence is critical for informing risk stratification, strengthening inpatient management, and guiding targeted interventions to reduce preventable deaths among PLHIV with cryptococcal meningitis in resource-limited settings.

## Methods

### Study design and setting

We conducted a retrospective cohort study using routinely collected medical records of adult PLHIV admitted with cryptococcal meningitis at KNRH in Kampala, Uganda, between January 2017 and December 2022. KNRH hosts the country’s largest tertiary infectious diseases unit and provides specialised inpatient care for HIV-related opportunistic infections, including cryptococcal meningitis and tuberculosis. As a national referral facility, KNRH receives patients from across Uganda, enhancing the relevance of findings for inpatient HIV care in similar resource-limited settings.

### Study population and eligibility criteria

The study population comprised adult patients admitted with HIV-related illnesses whose medical records documented a diagnosis of HIV-associated cryptococcal meningitis during the study period. Cryptococcal meningitis was defined by a positive cerebrospinal fluid CrAg test or India ink microscopy. Both ART-naive and ART-experienced patients were eligible, as were referral and non-referral cases.

Records were excluded if they lacked documented inpatient outcomes (e.g. missing discharge status), involved HIV-negative patients diagnosed with cryptococcal meningitis, or were unavailable for review. Medical records were retrieved and abstracted between June and September 2023.

### Sample size and sampling procedure

Sample size estimation followed the formula for unmatched cohort studies described by Kelsey et al. [16], assuming a two-sided significance level of 0.05 and 80% power. Mortality estimates were drawn from a comparable Tanzanian study reporting 14% mortality among HIV-positive patients without cryptococcal meningitis and 27% mortality among those with cryptococcal meningitis [17], yielding a minimum required sample size of 105 exposed patients.

In practice, a total population sampling approach was used. All available medical records of adult PLHIV admitted with cryptococcal meningitis during the study period and meeting the eligibility criteria were consecutively reviewed to maximise statistical power and representativeness. Of 3,042 HIV-related admissions, 650 cryptococcal meningitis cases were identified, and records for 634 patients (97.5%) were successfully retrieved and included in the final analysis.

### Data collection procedures

Data were abstracted from hard-copy medical records using a structured abstraction tool programmed in Kobo Collect, from 12^th^ December 2022 to 31^st^ January 2023. The tool was developed based on a review of published literature on predictors of cryptococcal meningitis mortality and relevant clinical guidelines. Variables captured included sociodemographic characteristics, clinical presentation, laboratory findings, treatment information, comorbidities, and in-hospital outcomes.

The abstraction tool underwent expert review by clinicians experienced in HIV and CM management and by epidemiologists familiar with retrospective cohort studies. Trained research assistants abstracted data using mobile devices, with daily supervisory checks for completeness and internal consistency. Identified discrepancies were resolved through cross-checking against original patient records.

### Data quality assurance

Data quality was strengthened through standardised abstraction procedures, training of research assistants, and routine supervisory review. During the pilot phase, internal consistency of the abstraction process was assessed and found to be acceptable. Content validity was ensured through expert review during tool development. Internal validity was further supported through consistent variable definitions and multivariable modelling to adjust for potential confounding.

### Data management and statistical analysis

Data were exported from Kobo Collect to Microsoft Excel and analysed using Stata version 15.Descriptive statistics summarised patient characteristics, with categorical variables presented as frequencies and percentages and continuous variables summarised using means (standard deviations) or medians (interquartile ranges), as appropriate.

Missing data were assessed for patterns and addressed using multiple imputation by chained equations, under the assumption that data were missing at random. Continuous variables were imputed using predictive mean matching, while categorical variables were imputed using logistic or multinomial regression. Multicollinearity among independent variables was assessed using variance inflation factors, with values greater than 10 considered indicative of problematic collinearity.

The proportion of cryptococcal meningitis among HIV-related admissions was calculated as the percentage of confirmed cases relative to all PLHIV admissions. In-hospital mortality was defined as death occurring during the admission in which cryptococcal meningitis was diagnosed.

Associations between explanatory variables and inpatient mortality were examined using logistic regression, as reliable time-to-event data were not consistently available to support survival analysis. Bivariate logistic regression was first performed, and variables with p ≤ 0.20 were included in a multivariable logistic regression model using a forward stepwise approach. Statistical significance in the multivariable model was set at p ≤ 0.05, and results are presented as adjusted odds ratios with 95% confidence intervals.

### Ethical considerations

Ethical approval was obtained from the Makerere University School of Public Health Higher Degrees Research and Ethics Committee (study protocol number 131), with a waiver of informed consent granted for retrospective review of de-identified medical records. Permission to access patient records was granted by KNRH. Data abstractors had no access to patient records that could identify them during and after data abstraction. All data were anonymised prior to analysis to ensure confidentiality.

## Results

### Patient admissions and records retrieval

Between January 2017 and December 2022, a total of 3,042 HIV-related inpatient admissions were recorded at KNRH in Uganda. Among these admissions, 650 patients (21.4%) were diagnosed with HIV-associated CM according to inpatient registers. Medical records for 634 of these patients (97.5%) were successfully retrieved and included in the analysis. Sixteen records (2.5%) could not be retrieved due to incomplete documentation and were excluded.

Of the 634 patients included, 249 died during hospitalization, corresponding to an overall in-hospital mortality of 39.3%. **Figure 1** summarises the selection and inclusion of study records.

**Figure 1:**
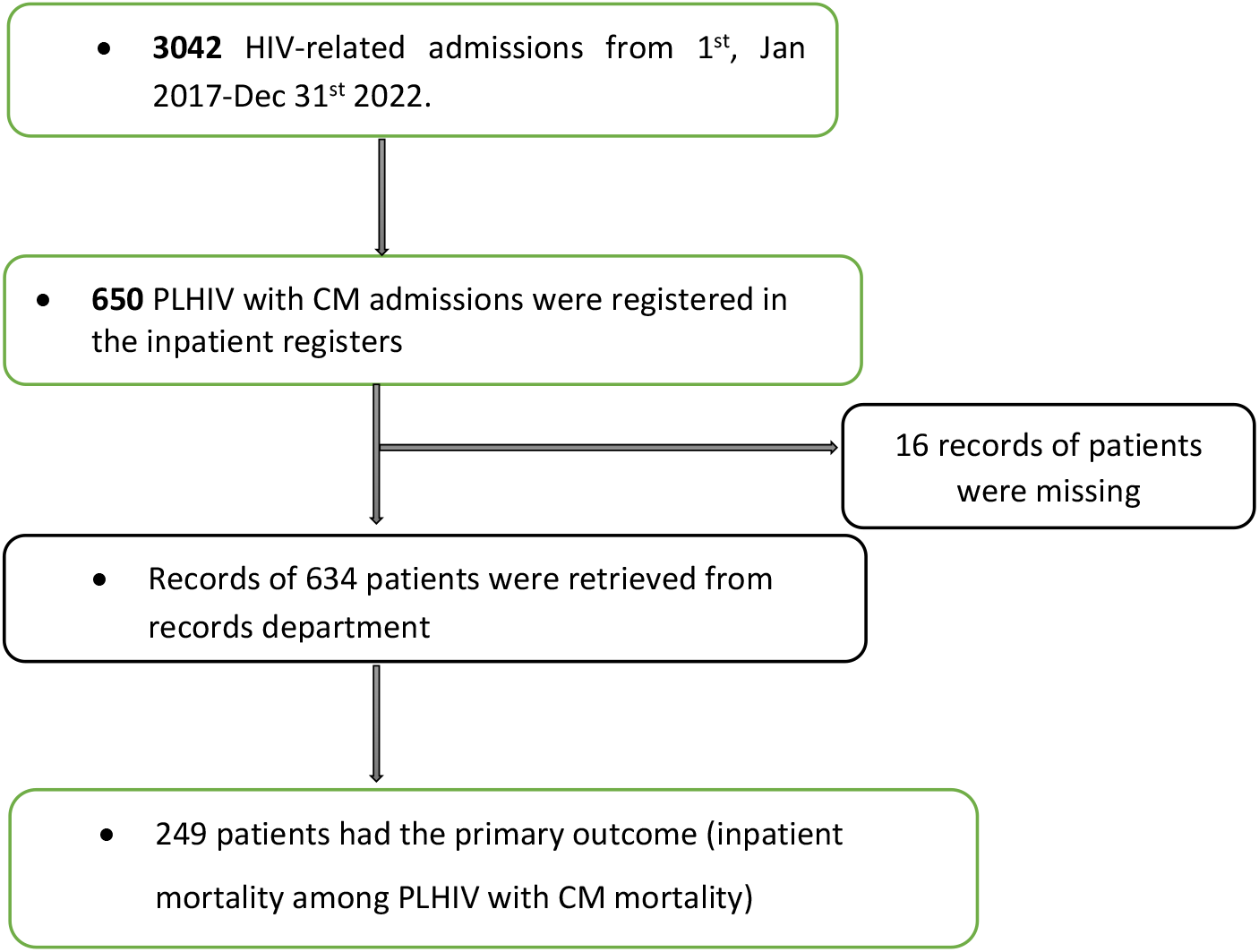
Medical records of adult PLHIV admitted for cryptococcal meningitis included the study.

### Socio-demographic and clinical characteristics

The median age of the study population was 36 years (interquartile range [IQR] = 15), with most patients aged 30–39 years (38.0%). Males accounted for 52.2% of participants. Duration of symptoms before admission was evenly distributed, with 31.1% presenting within one week, 37.4% within one to two weeks, and 31.5% after more than two weeks.

Headache was the most common presenting symptom (87.2%), followed by vomiting (58.7%) and blurring of vision (23.8%). A previous history of CM was documented in 35.0% of patients. The median length of hospital stay was 6 days (IQR = 10), with 50.3% of patients hospitalized for fewer than seven days.

Laboratory findings indicated advanced immunosuppression, with a median CD4 count of 45 cells/µL (IQR = 89). More than half of patients (52.5%) had CD4 counts below 50 cells/µL, and only 6.9% had CD4 counts above 200 cells/µL. Anaemia was common, with a mean haemoglobin level of 9.6 g/dL (SD = 3.3). Nearly half of patients (47.2%) had at least one concurrent opportunistic infection, most commonly tuberculosis, as shown in **Table 1**.

**Table 1:** Socio-demographic and clinical characteristics of the patients in the study.

| Parameter | Frequency<br>(N=634) | Percent<br>age (%) |
| --- | --- | --- |
| **Age (years) median= 36 (IQR=15) years** |  |  |
| <20 | 9 | 1.4 |
| 20-29 | 140 | 22.1 |
| 30-39 | 241 | 38.0 |
| 40-49 | 146 | 23.0 |
| 50-59 | 64 | 10.1 |
| 60+ | 34 | 5.4 |
| **Sex** |  |  |
| Male | 331 | 52.2 |
| Female | 303 | 47.8 |
| **Duration of hospitalization (days) median=6 (IQR=10) days** |  |  |
| <7 days | 319 | 50.3 |
| 7-14 days | 198 | 30.2 |
| >14 days | 117 | 18.5 |
| **Duration of symptoms before hospitalization** |  |  |
| Less than 1 week | 197 | 31.1 |
| 1-2 weeks | 237 | 37.4 |
| Greater than 2 weeks | 200 | 31.5 |
| **Duration of ART** |  |  |
| ART-naïve | 97 | 15.3 |
| Less than 1 year | 206 | 32.5 |
| 1-5 years | 223 | 35.2 |
| Greater than 5 years | 108 | 17.0 |
| **Previous history of cryptococcal meningitis** |  |  |
| Yes | 222 | 35.0 |
| No | 412 | 65.0 |
| **Signs, symptoms and comorbidities present at the time of diagnosis** |  |  |
| Headache | 553 | 87.2 |
| Vomiting | 372 | 58.7 |
| Blurring of vision | 151 | 23.8 |
| Cryptococcal meningitis IRIS | 73 | 11.5 |
| Concurrent Opportunistic infections (candidiasis, TB, e.tc) | 299 | 47.2 |
| Concurrent Diabetes mellitus (DM) | 30 | 4.7 |
| Concurrent cardiovascular disease (CVDs), hypertension (HTN), etc. | 26 | 4.1 |
| Concurrent CKD | 81 | 12.1 |
| **Hemoglobin level (g/dl) mean (SD)=9.6±(3.3)** |  |  |
| <8 | 223 | 35.2 |
| 8-10 | 163 | 25.7 |
| 10.1-12.5 | 109 | 17.2 |
| >12.5 | 139 | 21.9 |
| **CD4 T cell count (cell/uL) median CD4=45 (IQR=89) cells/uL** |  |  |
| <50 | 333 | 52.5 |
| 50-200 | 257 | 40.6 |
| >200 | 44 | 6.9 |

### Burden of cryptococcal meningitis and inpatient mortality

Cryptococcal meningitis accounted for 21.4% (650/3,042) of all HIV-related inpatient admissions during the study period. Among patients admitted with cryptococcal meningitis and included in the analysis, 39.3% (249/634) died during hospitalization.

### Factors associated with inpatient mortality among PLHIV with CM

At the bivariate level, several clinical and laboratory factors were associated with inpatient mortality at p ≤ 0.20 **(Table 2)**. Longer duration of hospitalization was associated with lower odds of death, with patients hospitalized for 7–14 days (crude odds ratio [COR] = 0.66; 95% CI: 0.46–0.95) and more than 14 days (COR = 0.57; 95% CI: 0.36–0.89) having reduced mortality compared with those hospitalized for fewer than seven days.

**Table 2:**
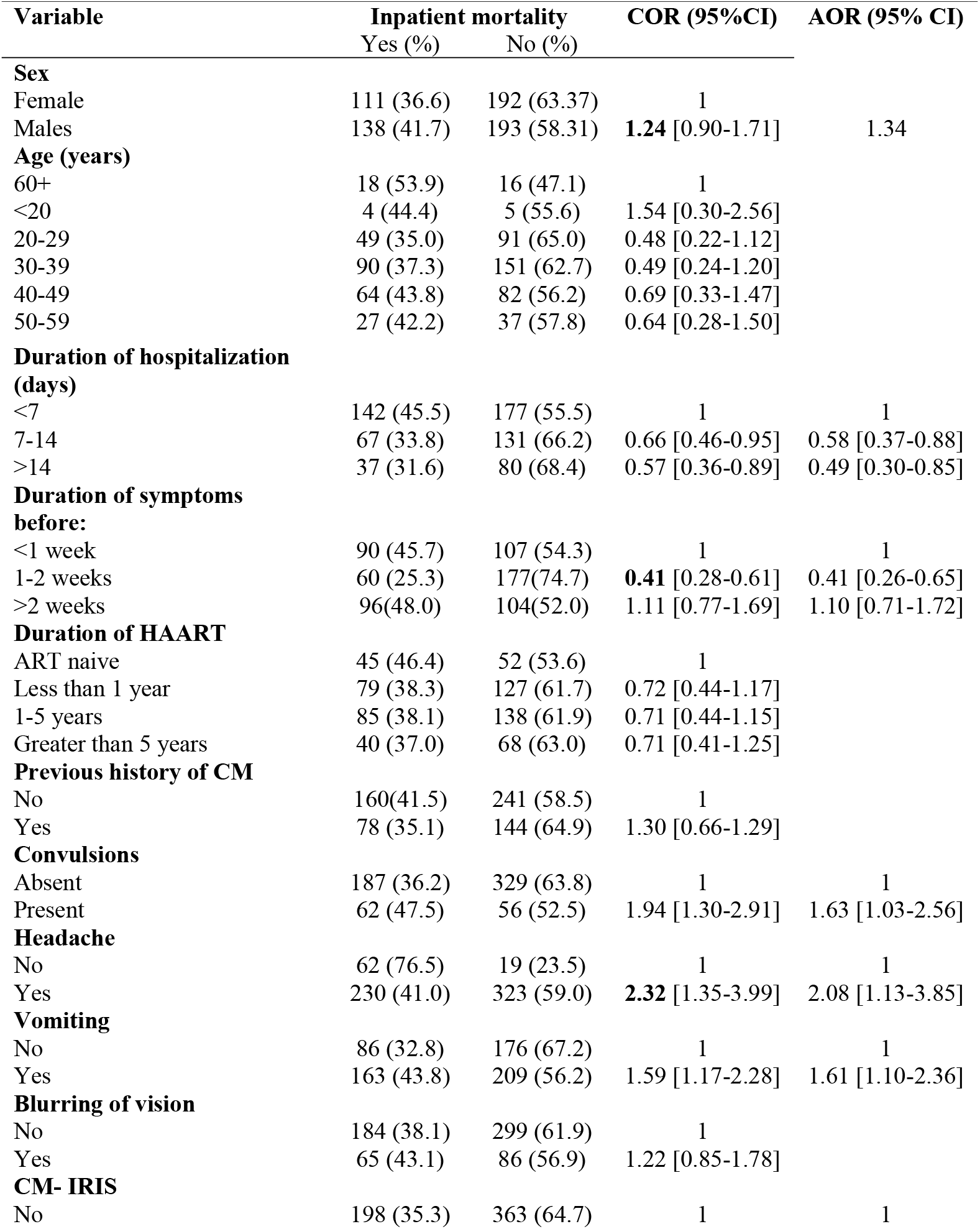
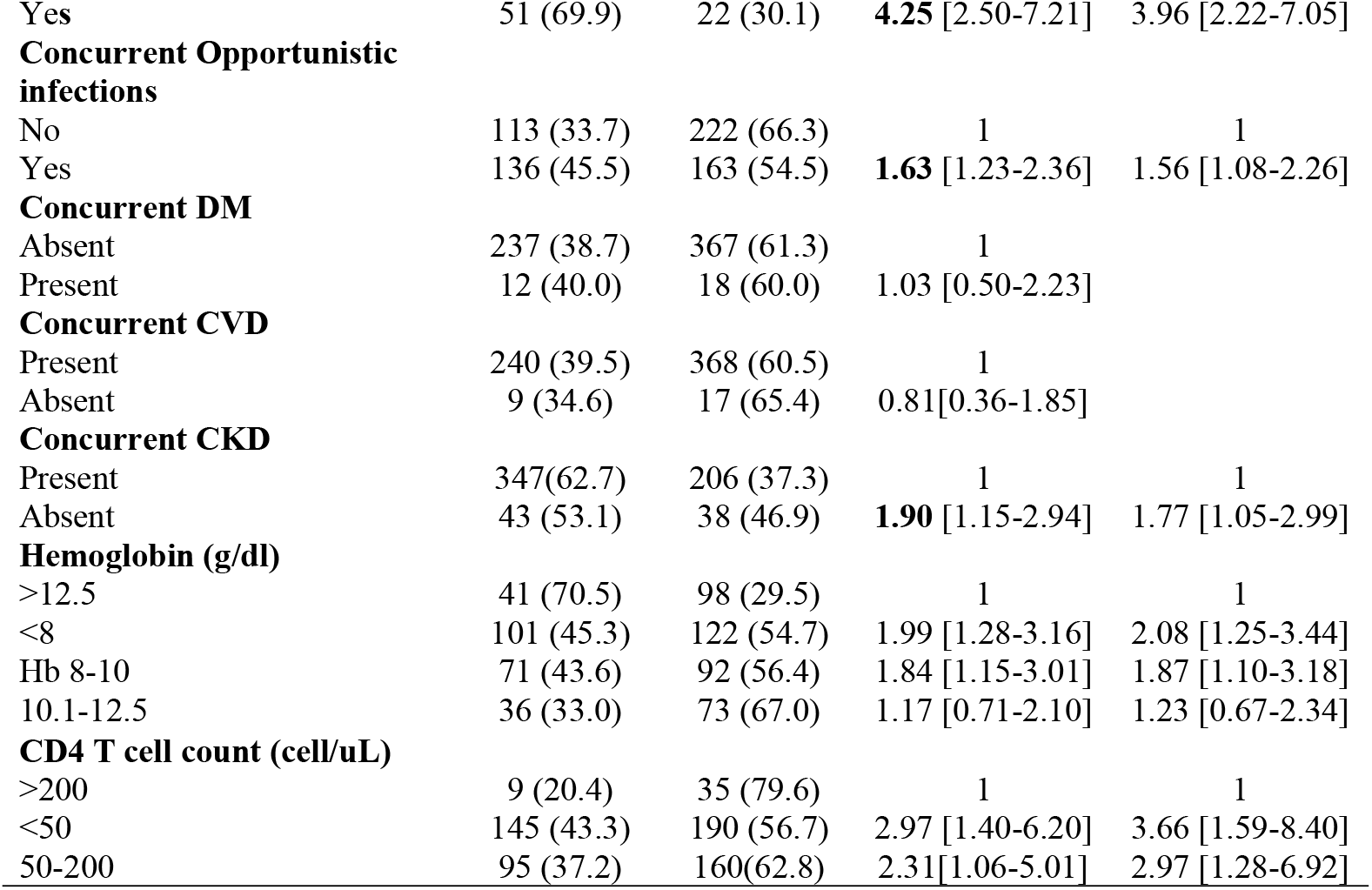
Clinical factors associated with inpatient mortality among PLHIV with CM at a national referral hospital in Uganda, between 2017 and 2022.

Symptom duration before admission of one to two weeks was associated with lower odds of death compared with presentation within one week (COR = 0.41; 95% CI: 0.28–0.61). Clinical features associated with higher mortality included convulsions, headache, vomiting, cryptococcal meningitis–associated immune reconstitution inflammatory syndrome (CM-IRIS), concurrent opportunistic infections, chronic kidney disease, anaemia, and low CD4 cell counts.

After adjustment for potential confounders, several factors remained independently associated with inpatient mortality. Longer hospitalization was associated with reduced odds of death, including stays of 7–14 days (adjusted odds ratio [AOR] = 0.58; 95% CI: 0.37–0.88) and more than 14 days (AOR = 0.49; 95% CI: 0.30–0.85). Presentation after one to two weeks of symptoms was also associated with lower mortality (AOR = 0.41; 95% CI: 0.26–0.65).

Clinical factors independently associated with increased odds of death included convulsions (AOR = 1.63; 95% CI: 1.03–2.56), headache (AOR = 2.08; 95% CI: 1.13–3.85), vomiting (AOR = 1.61; 95% CI: 1.10–2.36), CM-IRIS (AOR = 3.96; 95% CI: 2.22–7.05), concurrent opportunistic infections (AOR = 1.56; 95% CI: 1.08–2.26), and chronic kidney disease (AOR = 1.77; 95% CI: 1.05–2.99).

Anaemia remained strongly associated with mortality. Compared with patients with haemoglobin >12.5 g/dL, those with haemoglobin <8 g/dL (AOR = 2.08; 95% CI: 1.25–3.44) and 8–10 g/dL (AOR = 1.87; 95% CI: 1.10–3.18) had higher odds of death. Severe immunosuppression was also independently associated with mortality, with CD4 counts <50 cells/µL (AOR = 3.66; 95% CI: 1.59–8.40) and 50–200 cells/µL (AOR = 2.97; 95% CI: 1.28– 6.92) conferring higher risk compared with CD4 counts >200 cells/µL.

## Discussion

This study assessed the burden of inpatient mortality, and associated clinical factors among adult PLHIV with CM, admitted to a national referral hospital in Uganda between 2017 and 2022. CM meningitis accounted for more than one-fifth of HIV-related inpatient admissions, underscoring its continued contribution to morbidity among PLHIV despite expanded access to ART. This proportion is higher than pooled estimates reported in earlier systematic reviews, reflecting the persistent and disproportionate burden of cryptococcal disease in high-HIV-prevalence settings in SSA [7, 18].

In-hospital mortality among patients with CM was high (39.3%), consistent with reports from similar hospital-based studies across SSA [3, 15]. Most affected patients were young adults, highlighting the substantial loss of life among economically productive age groups. However, mortality was more strongly associated with clinical severity and comorbidity than with age alone, emphasising the importance of disease stage, immune status, and inpatient management in shaping outcomes [19].

Several clinical factors were independently associated with increased odds of inpatient mortality. Presenting symptoms such as headache, convulsions, and vomiting were common and likely reflect elevated intracranial pressure, a well-established predictor of poor outcomes in CM. Notably, intracranial pressure was documented in only a small proportion of patients, suggesting gaps in routine monitoring and adherence to guideline-recommended management in resource-limited inpatient settings [11]. Strengthening access to lumbar puncture equipment and training for pressure monitoring may therefore represent a critical opportunity to reduce preventable deaths.

In the present study, CM-IRIS was observed in approximately one in ten patients and was strongly associated with mortality. This finding aligns with previous studies from Uganda and other high-burden settings demonstrating elevated short-term mortality among patients who develop cryptococcal disease shortly after initiating ART [14, 20]. These results reinforce the importance of systematic cryptococcal antigen screening prior to ART initiation, careful timing of ART, and close clinical monitoring during early treatment to mitigate IRIS-related complications [11, 15].

Anaemia was highly prevalent and independently associated with mortality, with both moderate and severe anaemia conferring substantially increased risk of death. This finding is consistent with prior studies identifying anaemia as a marker of advanced disease, comorbidity, and poor physiological reserve among PLHIV with cryptococcal meningitis [13, 21]. Routine screening and timely management of anaemia should therefore be considered an integral component of inpatient care for CM.

Severe immunosuppression, reflected by low CD4 cell counts, and the presence of concurrent opportunistic infections were also associated with higher mortality. These findings underscore the compounded risk faced by patients presenting with advanced HIV disease and multiple coexisting conditions, which complicates clinical management and increases vulnerability to adverse outcomes [1, 3]. Earlier HIV diagnosis, linkage to care, and sustained viral suppression remain essential upstream strategies for reducing the burden and severity of CM.

Collectively, these findings highlight the need for strengthened health-system responses to cryptococcal meningitis in high-burden settings. Interventions should prioritise early case detection through point-of-care cryptococcal antigen screening, optimisation of ART timing, improved inpatient monitoring (including intracranial pressure assessment), and proactive management of anaemia and co-infections. Addressing these gaps has the potential to substantially reduce inpatient mortality among PLHIV with cryptococcal meningitis in resource-limited settings.

### Strengths and limitations

We analyzed a large, real-world dataset spanning six years from a national referral hospital in Uganda, enabling robust estimation of CM-related inpatient mortality, and associated clinical factors among PLHIV, under routine care conditions. However, several limitations should be considered. The retrospective design relied on routine medical records, which resulted in missing or incomplete documentation for some clinical variables, particularly intracranial pressure measurements. Additionally, the single-centre setting may limit generalisability to lower-level facilities, although the hospital’s national referral role enhances relevance for similar inpatient contexts. Finally, causal inferences cannot be drawn due to the observational nature of the study.

## Conclusion

Cryptococcal meningitis was associated with high in-hospital mortality among admitted adult PLHIV at a national referral hospital in Uganda between 2017 and 2022. Mortality correlated with identifiable clinical and health-system factors, including CM-IRIS, severe immunosuppression, anaemia, concurrent opportunistic infections, and markers of advanced disease at presentation.

Reducing mortality from HIV-associated cryptococcal meningitis requires strengthening early detection through routine cryptococcal antigen screening, optimising ART timing, improving inpatient monitoring and management, and addressing common comorbidities such as anaemia and renal disease. Further research is needed to evaluate context-appropriate treatment strategies and health-system interventions that can improve survival among PLHIV with cryptococcal meningitis in resource-limited settings.

## Data Availability

The dataset analyzed for this study is provided as supporting information (S1 File).

## Declarations

### Consent for publication

Not applicable

### Availability of data materials

The dataset analyzed for this study is provided as supporting information (S1 File).

## Acknowledgement

We acknowledge the staff at Kiruddu National Referral Hospital, Uganda for their assistance and guidance during data abstraction.

## Authors’ contributions

**Conceptualization:** Joshua Kitimbo^1*^, Esther Buregyeya, Noah Kiwanuka and Thomas Buyinza

**Methodology:** Joshua Kitimbo, Esther Buregyeya, Geofrey Mutole, John Paul Ibanda, Joseph Tumwine, Noah Kiwanuka, and Thomas Buyinza

**Formal analysis:** Joshua Kitimbo, Thomas Buyinza, Esther Buregyeya, Geofrey Mutole, John Paul Ibanda, Joseph Tumwine, and Noah Kiwanuka,

**Project administration:** Joshua Kitimbo

**Writing original draft:** Joshua Kitimbo

**Reviewing and editing final manuscript:** Joshua Kitimbo and Thomas Buyinza

## Supporting information

**S1 File. Data set**.

De-identified data set analyzed for this manuscript.

## References

1. Tugume L, Ssebambulidde K, Kasibante J, Ellis J, Wake RM, Gakuru J, et al. Cryptococcal meningitis. Nature Reviews Disease Primers. 2023;9(1):62.

2. McHale TC, Boulware DR, Kasibante J, Ssebambulidde K, Skipper CP, Abassi M. Diagnosis and management of cryptococcal meningitis in HIV-infected adults. Clinical microbiology reviews. 2023;36(4):e00156–22.

3. Williamson PR, Jarvis JN, Panackal AA, Fisher MC, Molloy SF, Loyse A, et al. Cryptococcal meningitis: epidemiology, immunology, diagnosis and therapy. Nature Reviews Neurology. 2017;13(1):13–24.

4. Muzazu SG, Assefa DG, Phiri C, Getinet T, Solomon S, Yismaw G, et al. Prevalence of cryptococcal meningitis among people living with human immuno-deficiency virus and predictors of mortality in adults on induction therapy in Africa: A systematic review and meta-analysis. Frontiers in Medicine. 2022;9:989265.

5. Mami D, Cuthrell KM, Manteghian M. Burden of HIV on Society Current Management Options and Future Prospective. International STD Research & Reviews. 2024;13(1):43–62.

6. Nkenfou C. Speaker presentations of the 2018 International Symposium on HIV and Emerging Infectious Diseases (ISHEID). Journal of Virus Eradication. 2018.

7. Rajasingham R, Govender NP, Jordan A, Loyse A, Shroufi A, Denning DW, et al. The global burden of HIV-associated cryptococcal infection in adults in 2020: a modelling analysis. The Lancet infectious diseases. 2022;22(12):1748–55.

8. Mohamed SH, Nyazika TK, Ssebambulidde K, Lionakis MS, Meya DB, Drummond RA. Fungal CNS infections in Africa: the neuroimmunology of cryptococcal meningitis. Frontiers in immunology. 2022;13:804674.

9. Okwir M, Link A, Rhein J, Obbo JS, Okello J, Nabongo B, et al., editors. High burden of cryptococcal meningitis among antiretroviral therapy–experienced human immunodeficiency virus–infected patients in Northern Uganda in the era of “test and treat”: implications for cryptococcal screening programs. Open forum infectious diseases; 2022: Oxford University Press US.

10. Iyer KR, Revie NM, Fu C, Robbins N, Cowen LE. Treatment strategies for cryptococcal infection: challenges, advances and future outlook. Nature Reviews Microbiology. 2021;19(7):454–66.

11. Alanazi AH, Adil MS, Lin X, Chastain DB, Henao-Martínez AF, Franco-Paredes C, et al. Elevated intracranial pressure in cryptococcal meningoencephalitis: examining old, new, and promising drug therapies. Pathogens. 2022;11(7):783.

12. Chakrabarti A, Patel AK, Soman R, Todi S. Overcoming clinical challenges in the management of invasive fungal infections in low-and middle-income countries (LMIC). Expert Review of Anti-infective Therapy. 2023;21(10):1057–70.

13. Tugume L, Morawski BM, Abassi M, Bahr NC, Kiggundu R, Nabeta HW, et al. Prognostic implications of baseline anaemia and changes in haemoglobin concentrations with amphotericin B therapy for cryptococcal meningitis. HIV medicine. 2017;18(1):13–20.

14. Bremer M, Kadernani YE, Wasserman S, Wilkinson RJ, Davis AG. Strategies for the diagnosis and management of meningitis in HIV-infected adults in resource limited settings. Expert Opinion on Pharmacotherapy. 2021;22(15):2053–70.

15. Rajasingham R, Smith RM, Park BJ, Jarvis JN, Govender NP, Chiller TM, et al. Global burden of disease of HIV-associated cryptococcal meningitis: an updated analysis. The Lancet infectious diseases. 2017;17(8):873–81.

16. Kelsey JL, Petitti DB, King AC. Key methodologic concepts and issues. See Ref. 1998;25:35–69.

17. Meda J, Kalluvya S, Downs JA, Chofle AA, Seni J, Kidenya B, et al. Cryptococcal meningitis management in Tanzania with strict schedule of serial lumber punctures using intravenous tubing sets: an operational research study. JAIDS Journal of Acquired Immune Deficiency Syndromes. 2014;66(2):e31–e6.

18. Nyazika T, Kamtchum-Tatuene J, Kenfak-Foguena A, Robertson VJ, Verweij PE, Meis JF, et al. Prevalence and mortality of cryptococcal meningitis in Africa from 1950 to 2017 and associated epidemiological mapping of C. neoformans and C. gattii species complexes: a systematic review and meta-analysis. C neoformans. 2019.

19. Stack M, Hiles J, Valinetz E, Gupta SK, Butt S, Schneider JG, editors. Cryptococcal meningitis in young, immunocompetent patients: a single-center retrospective case series and review of the literature. Open Forum Infectious Diseases; 2023: Oxford University Press US.

20. Howlett WP. Neurological disorders in HIV in Africa: a review. African health sciences. 2019;19(2):1953–77.

21. Yin S, Xu Y, Huang J, Xiong N, Han C, Ma K, et al. Immune reconstitution inflammatory syndrome secondary to autoimmune hemolytic anemia and cryptococcal meningitis. Frontiers in Neurology. 2019;10:812.

